# HbA1c and FIB-4 as Serologic Markers for the Risk of Progression of Stage A Heart Failure

**DOI:** 10.1101/2021.06.24.21259475

**Authors:** William Grigg, Faisal Mahfooz, Dharmista Chaudhary, Isain Zapata, Douglas Duffee

## Abstract

**Background:** The classic association of glycemic control as represented by glycosylated hemoglobin (Glyco% or HbA1c) with progression of micro and macro vascular clinical complications is well documented. However, use of the advanced glycation end product (AGE) axis as a marker for early diastolic hemodynamic changes leading to clinical heart failure has been suggested but is less well characterized. This study explored the association between elevated Glyco% and Fibrosis 4 (FIB-4, a 4-component marker for liver fibrosis) values and worsening measures of diastolic cardiac function in order to assess their utility as early serologic markers in cardiovascular disease prevention.

**Methods and Results:** A Retrospective cohort analysis was conducted in 102 patients presenting to the Parkview Medical Center health system who had received a full resting echo characterized by normal systolic ejection fraction and clinical risk factors associated with stage A heart failure in conjunction with Glyco% and FIB-4 scores all within a 3-month time window. Using regression analysis, measures of diastolic cardiac function were assessed in conjunction with rising Glyco% levels characterized as <6.5 and > 6.5 and FIB-4 scores after controlling for the presence of hypertension, coronary artery disease and valvular heart disease. Glyco% levels > 6.5 were significantly associated with a higher E/e’ ratio and closely associated with an elevated left atrial volume index both indicative of elevated left atrial pressure as a sensitive marker for diastolic cardiac dysfunction. FIB-4 scores did not appear to be clinically associated with progression of diastolic dysfunction.

**Conclusions:** Glyco%, long known to be a marker for metabolic glycemic control can also act as an early marker for identifying patients at increased risk for the progression of stage A heart failure. FIB-4 scores cannot.

**Registration:** https://clinicaltrials.gov/ct2/show/NCT04450576?term=duffee&draw=2&rank=3

## Introduction

The classic association of glycemic control, as represented by glyco%, with progression or improvement of microvascular and macrovascular clinical complications is well documented^1-5^. Additionally, while the association of glyco% elevation with an advanced clinical cardiovascular syndrome described as diabetic cardiomyopathy (beyond the classic cardiac risk factors associated with diabetes mellitus) has also been well documented the use of advanced glycation end products (AGE) as both a marker of and treatment target for the early diastolic hemodynamic changes of diabetic cardiomyopathy has been suggested but less well characterized^6-8,14^. In light of newer diabetic agents such as sglt2 inhibitors, which exert a cardio protective natriuretic effect in addition to a hypoglycemic effect, markers for early initiation of these medications both inside and outside the diabetic context, are important to elucidate. Stage A heart failure is defined as persons with normal cardiac structure and function who have predisposing risks for cardiovascular disease but have not yet manifested symptoms of heart failure^9-11^. Estimates suggest that Stage A heart failure may comprise greater than 50% of a community’s patient population and are patients commonly seen in primary care, endocrinology, cardiology and other clinics^5^.

In light of cardio metabolic agents such as sglt2 inhibitors which show promise in cardiovascular disease primary prevention^12-1^⍰, stage A heart failure patients may be an important target for a new therapeutic mechanism to deploy widely in preventative cardiovascular care. Showing an association between a commonly used metabolic marker and progressive hemodynamic cardiac disease would provide support for the logic of initiating medication therapy classically prescribed in the diabetic context to earlier use in the cardiac context with or without a concomitant indication for hypoglycemic therapy. Our study examined whether glyco% could be isolated as a hemodynamic marker of diastolic cardiac dysfunction in diabetics with stage A heart failure who may or may not yet need initiation or titration of hypoglycemic therapy. Secondarily a non-invasive easily obtainable marker for liver fibrosis (FIB-4) was also assessed for its correlation to diastolic function and potential utility as a treatment marker.

## Methods

### Participants

A community-based population cohort registry of all patients was identified by retrospective chart review who had presented to Parkview Medical Center’s in-patient and outpatient services in Pueblo County Colorado from 1/2019 to 6/2020 and who had obtained a full resting echo characterized by normal LV systolic function in conjunction with glyco%, brain natriuretic peptide (BNP), CBC and CMP within a concomitant 3-month time frame. 2263 patients were identified. A sample of 102 participants was randomly selected for this study. The selected sample size was determined by power analysis (1-β>0.8) for detection of a 5% change in Left atrial volume (LAV) examining the registry from most to less recent index encounter to a p value less than 0.05 until the N value was met. We also included a measure of liver fibrosis as estimated by the FIB-4 equation to assess for an association between a non-invasive marker for liver fibrosis and the same measures of diastolic cardiac function and a measure of the glycemic status of study groups at a cutoff level of 6.5 to include those with mild diabetes who may not yet require hypoglycemic therapy according to current guidelines.^15^ Patient’s age, gender (M=Male, F=Female) Body Mass index (BMI), Body Surface Area (BSA), Brain Natriuretic Peptide (BNP), previous history of hypertension (HTN), cardiac valvular disease and Coronary Artery Disease (CAD) were also recorded. All data was compiled from retrospective medical records for the participants selected for the sample. This study was sanctioned and approved by the Parker Medical Center Institutional Review Board.

### Echocardiography

Transthoracic echocardiography was performed in accordance with published standards and interpreted by a board-certified cardiologist independent of any knowledge of this study^16^. The presence of hypertension was defined as greater than 140/90 measured closest in time to the index echocardiogram. Valvular disease was defined as any regurgitation or stenosis noted to be greater than mild. Coronary artery disease was defined as any of the following: any regional wall motion abnormality on echo and/or cardiac catheterization showing at least one vessel with 50% or more stenosis. All labs were processed through Parkview’s central laboratory.

Statistical analysis--Means, standard deviation and range were calculated for continuous variables while frequencies and proportional percentages were calculated for categorical variables. All descriptive statistics were calculated using PROC MEANS and PROC FREQ from SAS/STAT v.9.4 (SAS Institute, Cary NC). Generalized Additive Models (GAMs) were used to evaluate the association of the dependent variables Glycemic percentage (Glyco%) and liver Fibrosis (Fib-4) to the independent variables of HTN, Valvular disease, CAD, Age, BMI, BNP, LAVI and E/e’. Effects were introduced into a semiparametric model by including the effects of gender, HTN, valve disease and CAD as parametric independent variables (categorical variables) and age, BMI, BNP, LAVI and E/e’ as smoothing splines with 3 degrees of freedom (continuous variables). Models were fitted independently by dependent variable (Glyco% and Fib-4) and assumed a Gaussian distribution for the residuals. Some variables were removed from the final model to avoid multi-collinearity: first, BMI and BSA which are intimately correlated^17^ and thus, BMI was the only one included; second, E, Lateral e’ and E/e’ are functionally related with each other and thus only E/e’ was included in the model; last, LAVI and LADI where only LAVI was included in the model since it is the strongest method to predict any and moderate to severe diastolic dysfunction. ^18^ All modeling was performed using PROC GAM in SAS/STAT v.9.4. Modeling effects are displayed as Risk Ratios with their 95% confidence intervals and non-linearity associations of the continuous variables evaluated as smoothing splines are evaluated through an analysis of deviance using a cubic pattern (3 degrees of freedom). Smoothing component plots included a 95% curve-wise Bayesian confidence band for each component.. Two-tailed P-values <0.05 were considered statistically significant

## Results

In total, 102 patients were retrospectively identified who met inclusion and exclusion criteria. Their descriptive characteristics which include mean, standard deviation and range of values are shown in Table 1 for continuous variables and frequency and percentage proportion are shown in Table 2 for categorical variables. Patients with a left ventricular ejection fraction less than 50% were excluded. Parameter estimates obtained through GAMs displayed as risk ratio significant association (p< 0.05) between Glyco% (HbA1c) with BMI (P=0.0043) and E/e’ (P=0.0345). For both models, Glyco% and Fib-4, neither sex nor comorbidity confounders of CAD, Valve disease and HTN were significantly associated. No significant associations to Fib-4 in general were detected in this model mode. However, spline smoothing components for linearly defined effects are presented in Figure 2 where only one significant spline smoothing component was detected for BNP (P=0.0019, through analysis of deviance) in the Fib-4 model. Although its association remained as non-significant towards Fib-4 prediction (P=0.2043).

**Table 1:** Descriptive statistics continuous parameters evaluated in the study cohort

| Variable | Mean | Std Dev | Range |
| --- | --- | --- | --- |
| **Age** <sub>yrs</sub> | 69.245 | 12.916 | (31 - 94) |
| **BMI** <sub>kg/m2</sub> | 30.559 | 9.055 | (14.8 - 55) |
| **BSA** <sub>m2</sub> | 1.941 | 0.334 | (1.28 - 2.85) |
| **Glyco%** | 6.850 | 1.879 | (4.7 - 14) |
| **BNP** <sub>pg/ml</sub> | 2487.9 | 4733.9 | (6 - 35000) |
| **Fib4** | 2.427 | 2.773 | (0.26 - 21.47) |
| **LAVI** <sub>ml/m2</sub> | 34.176 | 12.372 | (11 - 69) |
| **LADI** <sub>cm/m2</sub> | 2.077 | 0.473 | (1.2 - 4.1) |
| **E** <sub>m/sec</sub> | 1.005 | 0.346 | (0.088 - 2.074) |
| **Lat e'** <sub>m/sec</sub> | 0.107 | 0.161 | (0.035 - 1.23) |
| **E/e'** | 13.936 | 6.041 | (4 - 34) |

**Table 2:** Descriptive statistics of categorical parameters evaluated in the study cohort

| **Variable** | **Frequency** | **Percent** |
| --- | --- | --- |
| **Gender** |  |  |
| **Female** | 58 | 56.86 |
| **Male** | 44 | 43.14 |
| **CADrwma** |  |  |
| **No** | 76 | 74.51 |
| **Yes** | 26 | 25.49 |
| **Valve Disease** |  |  |
| **No** | 95 | 93.14 |
| **Yes** | 7 | 6.86 |
| **HTN** |  |  |
| **No** | 23 | 22.55 |
| **Yes** | 79 | 77.45 |

**Figure 1:**
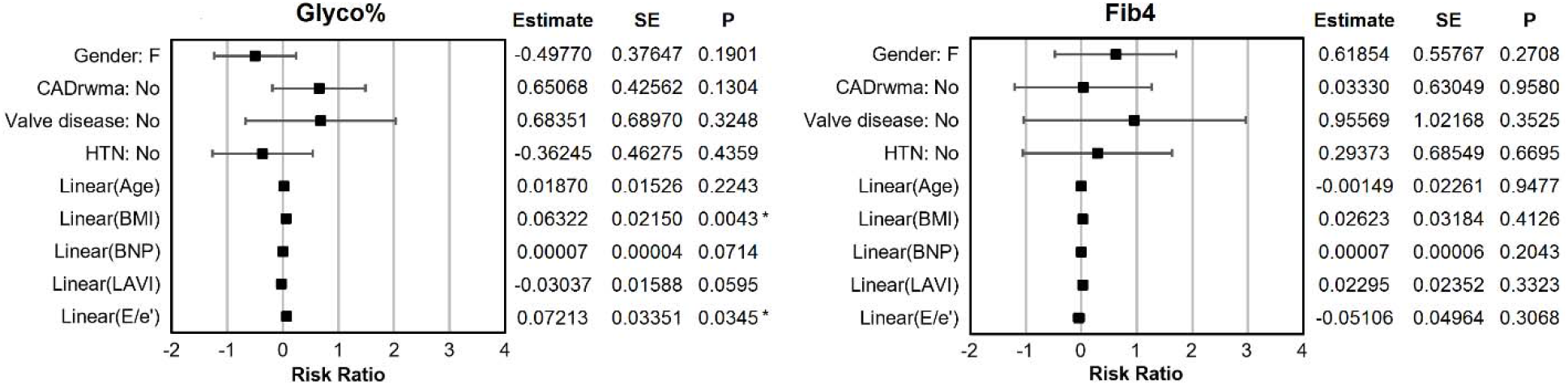
Risk ratio parameter estimates of Glyco% and Fib-4 models. Sex (M/F) and comorbidities (Yes/No) were defined as parametric responses while age, BMI, BNP, LAVI and E/e’ were defined as spline smoothing components. Significantly associated effects (P<0.05) are marked by an asterisk (*).

**Figure 2:**
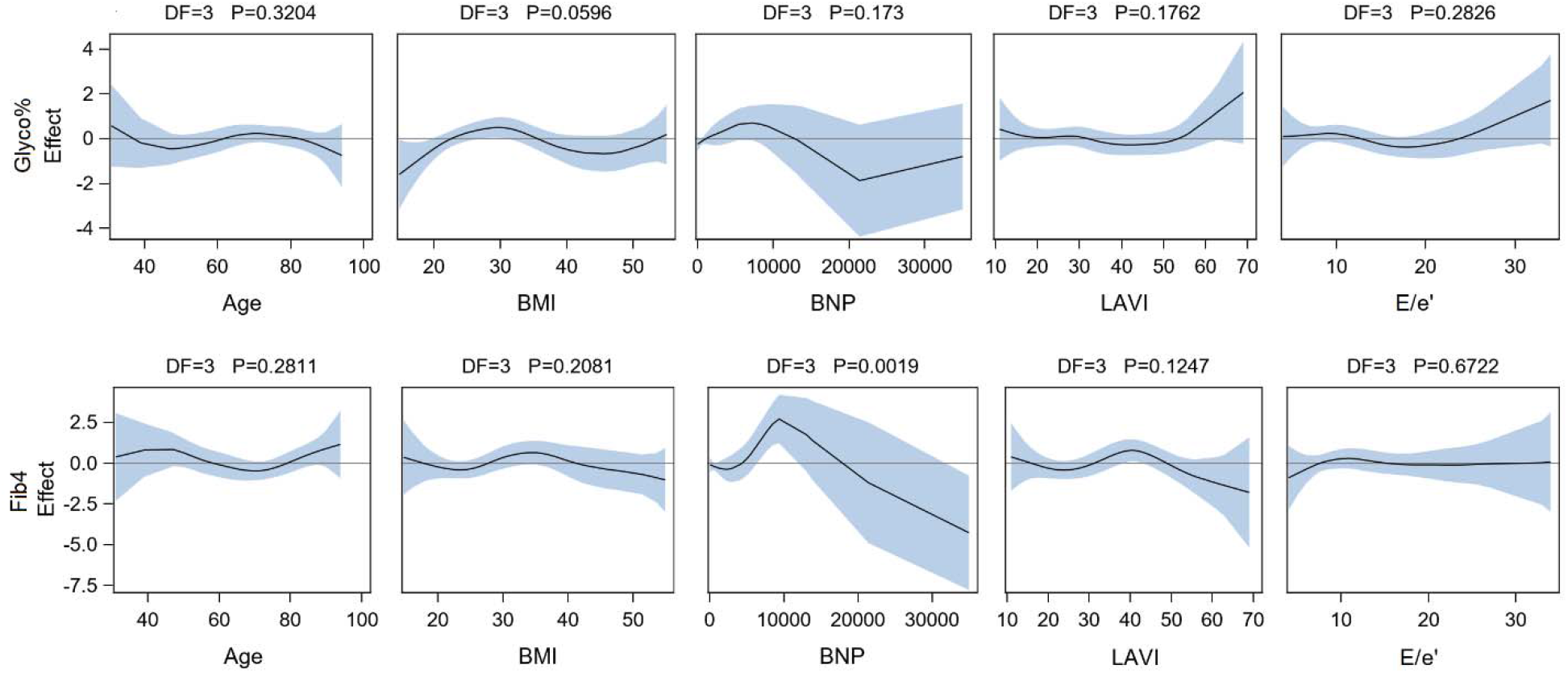
Spline smoothing component evaluation of deviance for linearly defined parameters in the generalized additive models. Curve-wise confidence band is at 95% confidence level. P values presented are for the analysis of deviance of each spline smoothing component where significant deviance is declared at P<0.05.

On a subsequent model iteration where parameter estimates were differentiated based on Glyco% grouping low group vs high group (the “low group” being defined as having Glyco% 6.5 or less while the “high group” being defined as having Glyco% over 6.5) detected only increased risk associated to age for the low group when estimating Glyco%. This data is presented in Table 3. Also in these models, neither sex nor comorbidity associations to Glyco% and Fib-4 were detected. In summary, some predictive potential for BMI and E/e’ was observed for Glyco% models adjusted for comorbidities although no comorbidities were associated. For low group vs high group models only age was detected as significantly predictive. No predictive potential was detected in general for Fib-4 criteria in the main model and in the subsequent low group vs high group model iteration.

**Table 3:**
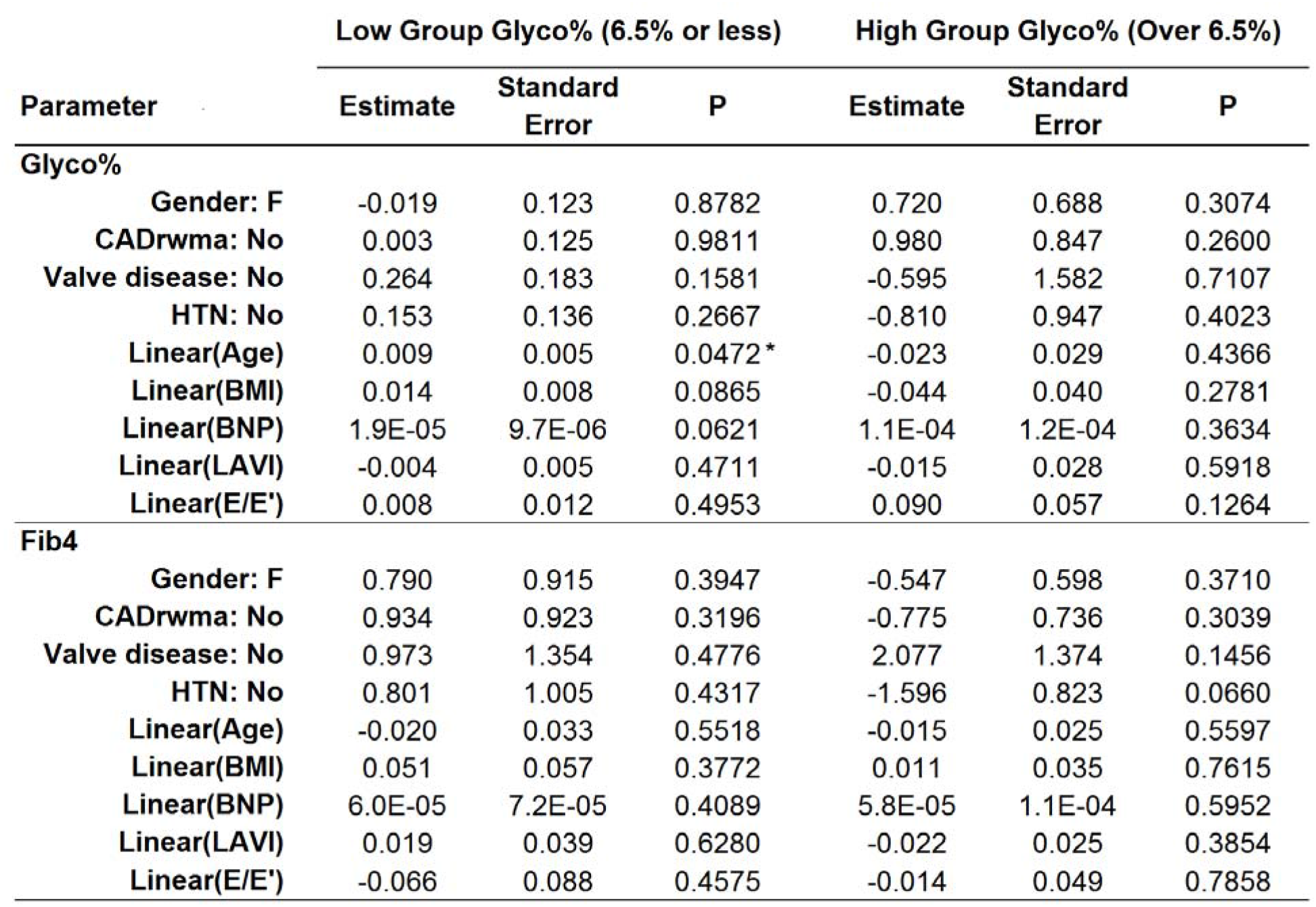
Low group vs high group parameter estimates. “Low group” being defined as having Glyco% 6.5 or less while the “high group” being defined as having Glyco% over 6.5. Groupings were evaluated independently for each Glyco% and Fib-4. Sex (M/F) and comorbidities (Yes/No) were defined as parametric responses while age, BMI,BNP, LAVI and E/e’ were defined as spline smoothing components. Significantly associated effects (P<0.05) are marked by an asterisk (*).

## Discussion

AGE formation is characterized by the glycation of plasma proteins during longstanding hyperglycemia. These AGE’s then stimulate transmembrane receptors of advance glycation end products (rAGE) on somatic cells including cardiac myocytes resulting in alteration of intracellular signaling, gene expression, release of pro-inflammatory molecules and free radicals. ^19^ These processes at least in part are thought to play a role in the development of cardiac fibrosis independent of the classic vascular complications related to diabetes mellitus. In contrast to transmembrane rAGE, soluble receptors for advance glycation end products (sRAGE) function to reduce risk associated with transmembrane rAGE by the pre-transmembrane binding of AGE’s and subsequent reduction of intracellular rAGE signaling. Specific sRage’s have been identified and may have the potential to serve as markers for AGE related cardiac risk. ^20^ As an early glycation end product, glyco% is nicely positioned as a hemodynamic risk marker inside this mechanism. Additionally, as lipids are thought to play a role in hepatic steatosis and liver fibrosis as well as in cross linking to form more advanced glycation end products capable of stimulating transmembrane rAGE, easily obtainable markers for liver fibrosis could serve as markers for cardiac fibrosis^21^ which is what prompted the evaluation of FIB-4 in this study.

Stage A heart failure patients, namely those with preclinical risk factors for progression to more advanced heart failure stages are an attractive cohort to study in that early intervention may provide significant attenuation of disease in this patient population. The definition of stage A heart failure in this study was applied to a cohort primarily defined by normal LV systolic function. In controlling for patients with underlying hypertension, CAD and valvular heart disease we further isolated the cohort to a stage A definition. ^22^ The difficulty was in using diastolic dysfunction as an independent variable which could be seen as excluding our cohort from true stage A characteristics. In that the degree of diastolic dysfunction based on E/A changes needed to advance patients from the stage A category is not clear in the definition of stage A heart failure, we tried to utilize the more sensitive “pre-clinical” markers for left atrial pressure and chronic diastolic dysfunction LAVI and E/e’.

The use of the dependent variables glyco% and FIB-4 was designed to capture simple and easily obtainable markers for cardiac fibrosis. In that current diabetes guidelines recommend a goal for hypoglycemic therapy to a glyco% less than 7.5, defining the study group as those who meet a definition for DM but may not yet require hypoglycemic therapy makes the results more applicable for cardiac risk reduction as opposed to simple glycemic control. FIB-4, used as a marker for liver fibrosis which is often associated with hyperglycemia and hyperlipidemia and of the same potential mechanism as metabolic cardiac fibrosis was attractive in that age as detected in our low group model iteration, platelets, AST and ALT are its formulaic components, however we did not find an association between FIB-4 and measures of diastolic dysfunction.

2 measures of diastolic function were used to assess the AGE-myocardial hypertrophy hypothesis. ^23^ Left atrial volume was seen as reflective of “the cumulative chronic effect of LV filling pressure over time”. The E/e’ ratio was also used as a doppler measure of diastolic function because it combines the ratio of a trans mitral flow measurement (a marker for the trans mitral pressure gradient) with a tissue doppler measurement (a marker of LV diastolic pressure) which is more sensitive for diastolic dysfunction. ^24^

Distinguishing glyco% as a compliance marker for progressive cardiac diastolic dysfunction beyond a non-specific vascular risk marker has been demonstrated. ^25^ This distinction is what sets diabetic cardiomyopathy apart from standard diabetic vascular risk factors and raises the specter of earlier hemodynamic primary preventative cardiac therapy even earlier then standard glycemic goals may dictate, especially in light of sglt2 inhibitor effects.

## Conclusion

In patients with clinical cardiovascular risk characterized as stage A heart failure, glyco% can be used as a sensitive marker to identify those most at risk for disease progression based on diastolic hemodynamic parameters. Additionally, the glyco% can be used to identify those who may benefit from preventative cardiovascular treatment via the natriuretic effects of newer glycemic agents prior to a glycemic indication for therapy.

## Supporting information

Strobe Checklist

## Data Availability

Data is stored in Parkview Medical Center secure drive

