## Supplementary material for "HbA1c and FIB-4 as Serologic Markers for the Risk of Progression of Stage A Heart Failure": Strobe Checklist

STROBE Statement—Checklist of items that should be included in reports of ***cohort studies***

Title Page

First author: Grigg DO

Authors:

William Grigg DO 1, Faisal Mahfooz MD 1, Dharmista Chaudhary MD 1, Isain Zapata PhD 2, Douglas Duffee MD, MDiv, FACP 1, 3.

Affiliations:

1 Department of Internal Medicine, Parkview Medical Center, Pueblo, CO 81003

2 Department of Biomedical Sciences, Rocky Vista University, Parker, CO 80134

3 Department of Graduate Medical Education, Parkview Medical Center, Pueblo, CO 81003

NO organizational funding support

Corresponding author:

Douglas Duffee MD, MDiv, FACP. Mail Address: 315 W. 15th St. Pueblo, Colorado 81003. Phone: +1(719)595-7968.

Total word count = 3200

Total pages = 15

Subject terms: Diabetic Cardiomyopathy, HbA1c, Glyco%, Fib-4 index, Diastolic dysfunction, Stage A heart failure

Abstract

Background-- The classic association of glycemic control as represented by glycosylated hemoglobin (Glyco% or HbA1c) with progression of micro and macro vascular clinical complications is well documented. However, use of the advanced glycation end product (AGE) axis as a marker for early diastolic hemodynamic changes leading to clinical heart failure has been suggested but is less well characterized. This study explored the association between elevated Glyco% and Fibrosis 4 (FIB-4, a 4-component marker for liver fibrosis) values and worsening measures of diastolic cardiac function in order to assess their utility as early serologic markers in cardiovascular disease prevention.

Methods and Results-- A Retrospective cohort analysis was conducted in 102 patients presenting to the Parkview Medical Center health system who had received a full resting echo characterized by normal systolic ejection fraction and clinical risk factors associated with stage A heart failure in conjunction with Glyco% and FIB-4 scores all within a 3-month time window. Using regression analysis, measures of diastolic cardiac function were assessed in conjunction with rising Glyco% levels characterized as <6.5 and > 6.5 and FIB-4 scores after controlling for the presence of hypertension, coronary artery disease and valvular heart disease. Glyco% levels > 6.5 were significantly associated with a higher E/e’ ratio and closely associated with an elevated left atrial volume index both indicative of elevated left atrial pressure as a sensitive marker for diastolic cardiac dysfunction. FIB-4 scores did not appear to be clinically associated with progression of diastolic dysfunction.

Conclusions—Glyco%, long known to be a marker for metabolic glycemic control can also act as an early marker for identifying patients at increased risk for the progression of stage A heart failure. FIB-4 scores cannot.

|  | Item No | Recommendation | Page No |
| --- | --- | --- | --- |
| **Title and abstract** | 1 | (*a*) Indicate the study’s design with a commonly used term in the title or the abstract |  |
|  |  | (*b*) Provide in the abstract an informative and balanced summary of what was done and what was found | 1-2 |
| Introduction | | | |
| Background/rationale | 2 | Explain the scientific background and rationale for the investigation being reported | 2 |
| Objectives | 3 | State specific objectives, including any prespecified hypotheses | 2 |
| Methods | | | |
| Study design | 4 | Present key elements of study design early in the paper | 4 |
| Setting | 5 | Describe the setting, locations, and relevant dates, including periods of recruitment, exposure, follow-up, and data collection | 4 |
| Participants | 6 | (*a*) Give the eligibility criteria, and the sources and methods of selection of participants. Describe methods of follow-up | 4 |
|  |  | (*b*) For matched studies, give matching criteria and number of exposed and unexposed |  |
| Variables | 7 | Clearly define all outcomes, exposures, predictors, potential confounders, and effect modifiers. Give diagnostic criteria, if applicable | 4 |
| Data sources/ measurement | 8* | For each variable of interest, give sources of data and details of methods of assessment (measurement). Describe comparability of assessment methods if there is more than one group | 4 |
| Bias | 9 | Describe any efforts to address potential sources of bias | 7 |
| Study size | 10 | Explain how the study size was arrived at | 4 |
| Quantitative variables | 11 | Explain how quantitative variables were handled in the analyses. If applicable, describe which groupings were chosen and why | 5 |
| Statistical methods | 12 | (*a*) Describe all statistical methods, including those used to control for confounding | 5 |
|  |  | (*b*) Describe any methods used to examine subgroups and interactions |  |
|  |  | (*c*) Explain how missing data were addressed |  |
|  |  | (*d*) If applicable, explain how loss to follow-up was addressed |  |
|  |  | (*e*) Describe any sensitivity analyses |  |
| Results | | | 5-6 |
| Participants | 13* | (a) Report numbers of individuals at each stage of study—eg numbers potentially eligible, examined for eligibility, confirmed eligible, included in the study, completing follow-up, and analysed | 4 |
|  |  | (b) Give reasons for non-participation at each stage |  |
|  |  | (c) Consider use of a flow diagram |  |
| Descriptive data | 14* | (a) Give characteristics of study participants (eg demographic, clinical, social) and information on exposures and potential confounders | 4 |
|  |  | (b) Indicate number of participants with missing data for each variable of interest |  |
|  |  | (c) Summarise follow-up time (eg, average and total amount) |  |
| Outcome data | 15* | Report numbers of outcome events or summary measures over time | 5-6 |

| Main results | 16 | (*a*) Give unadjusted estimates and, if applicable, confounder-adjusted estimates and their precision (eg, 95% confidence interval). Make clear which confounders were adjusted for and why they were included | 5-6 |
| --- | --- | --- | --- |
|  |  | (*b*) Report category boundaries when continuous variables were categorized |  |
|  |  | (*c*) If relevant, consider translating estimates of relative risk into absolute risk for a meaningful time period |  |
| Other analyses | 17 | Report other analyses done—eg analyses of subgroups and interactions, and sensitivity analyses | 5 |
| Discussion | | | |
| Key results | 18 | Summarise key results with reference to study objectives | 5-6 |
| Limitations | 19 | Discuss limitations of the study, taking into account sources of potential bias or imprecision. Discuss both direction and magnitude of any potential bias | 7 |
| Interpretation | 20 | Give a cautious overall interpretation of results considering objectives, limitations, multiplicity of analyses, results from similar studies, and other relevant evidence | 7-8 |
| Generalisability | 21 | Discuss the generalisability (external validity) of the study results | 8 |
| Other information | | | |
| Funding | 22 | Give the source of funding and the role of the funders for the present study and, if applicable, for the original study on which the present article is based | 1 |

*Give information separately for exposed and unexposed groups.

**Note:** An Explanation and Elaboration article discusses each checklist item and gives methodological background and published examples of transparent reporting. The STROBE checklist is best used in conjunction with this article (freely available on the Web sites of PLoS Medicine at http://www.plosmedicine.org/, Annals of Internal Medicine at http://www.annals.org/, and Epidemiology at http://www.epidem.com/). Information on the STROBE Initiative is available at http://www.strobe-statement.org.
